# Failure of *R*_0_ near the epidemic threshold in the classical SIS model

**DOI:** 10.1101/2023.09.18.23295738

**Authors:** Mozzamil Mohammed

## Abstract

The primary predictor of a disease outbreak and severity is the basic reproduction number *R*_0_, which represents the average number of secondary cases produced by introducing an infected individual into an entirely susceptible population. According to the classical SIS model, a disease with *R*_0_ less than one will eventually die out and persist if *R*_0_ is greater than one. Using the pair-approximation method, we reconstruct the classical SIS model by explicitly accounting for neighbourhood interactions between susceptible and infected individuals. Specifically, the disease can only be transmitted, with some transmission probability, if a susceptible individual is surrounded by at least one infected individual within its direct neighborhoods. Despite the simplicity of the SIS model present here, results produced by the pair-approximation model deviates significantly from predictions by the mean-field approximation model, particularly near the epidemic threshold. Contrasting the standard SIS model based on the mean-field approach, we find scenarios where the disease dies out even if *R*_0_ is greater than one. We suggest a crucial need for redefining the basic reproduction number on a smaller spatial scale and taking the average *R*_0_ over a global scale, rather than applying it globally to an entire population. However, in the realm of more intricate models of infectious diseases, it remains an open question to what extent mean-field approximation predictions diverge from predictions produced by models that consider neighborhood interactions.

## 1 Introduction

The basic reproduction number *R*_0_ quantifies the average number of secondary infections produced by an infected individual when introduced into a completely susceptible population (Dietz, 1993). The basic reproduction number has long been considered as import predictor of a disease spread and severity, and often plays an important role for supporting decisions of public health officials and policymakers (Guerra et al., 2017; Zhao et al., 2020). Estimation of the basic reproduction number is typically complex as it depends on various biological and environmental factors (Delamater et al., 2019). However, in the simple classical susceptible-infected-susceptible (SIS) model, the basic reproduction number emerges from just two key parameters and is defined as the average disease transmission rate divided by average recovery rate per an infected individual (Allen, 1994). It is commonly accepted that a disease with *R*_0_ less than 1 will eventually die out and persist if *R*_0_ is greater than 1. The epidemic threshold, occurring at *R*_0_ equal to 1, represents the boundary between the extinction and sustained transmission of the disease. The outcome of a disease transmission away from the epidemic threshold might be intuitively straightforward, depending on the transmission complexity.

However, estimation of *R*_0_ for the same disease can vary as the model set up and complexity change. Depending on the method used to calculate *R*_0_, many diseases with an *R*_0_ value greater than 1 are predicted to die out and persist when *R*_0_ is less than 1, leading to a failure of *R*_0_ (Li et al., 2011). The basic reproduction number is primarily defined on a global scale, often disregarding the small local scale on which interactions between susceptible and infected individuals occur. In reality, an infected individual cannot infect a susceptible one if they are spatially distant. Nevertheless, it is commonly assumed by the mean-field approximation method that all individuals within a population do interact with one another, and this simplification has been widely accepted. Network and pair-approximation models of infectious diseases typically account for interactions between neighbouring pairs of individuals (Keeling, 1999; Keeling and Eames, 2005; Payne, 2019). Such models often describe the dynamics of infectious disease more accurately than the mean-field approximation models. The pair-approximation method has been employed to describe systems other than infectious diseases where neighborhood interactions are crucial determinants of the dynamics outcomes (Harada and Iwasa, 1994; Mohammed et al., 2023, 2018). This particular methodology correctly captures systems dynamics predicted lattice-based models, offering mathematical tractability.

Using the pair-approximation method, we reconstruct the classical SIS model by explicitly accounting for neighborhood interactions among susceptible and infected individuals. The pair-approximation approach assumes that a susceptible individual can only get infected if it is surrounded by at least one infected individual present in its direct neighbourhoods. That is a necessary condition for a disease transmission, but the transmission occurs with some probability, following an interaction between susceptible and infected individuals. The core object of this work is to assess whether the *R*_0_ concept applies when neighbourhood interactions are taken into account in infectious disease models. Although the SIS model presented here is simple, but results of the pair-approximation model deviates significantly from previous predictions by the mean-field approximation model. Contrasting the SIS model that based mean-field approximation, the pair-approximation model exhibits a scenario where a disease can die out even if *R*_0_ is greater than 1. This divergence in the predictions of the two models occurs mainly around the epidemic threshold. More complex models of infection diseases based on the mean-field approximation are thus expected to diverge more significantly from models that explicitly account for neighbourhood interactions. We suggest that estimations of the basic reproduction number should be redefined on a small neighbourhood scale, rather than on a global scale for the entire population. Estimation of the basic reproduction number might not be enough to inform public health officials about the current and future behaviour of the disease outbreak dynamics, and this fundamental concept of *R*_0_ can be misleading by omitting some of its important properties such as spatial scales.

## 2 Methods

### 2.1 Pair-approximation model of disease transmission

Pair approximation is a method of constructing ordinary differential equation models, describing the dynamics of neighbouring pairs, the global and local dynamics of a given population (Harada and Iwasa, 1994). It provides an explicit description for neighborhood interactions among individuals and accurately capture the spatiotemporal dynamics predicted by lattice-based models (Mohammed et al., 2023).

Here, we use the pair-approximation method to reconstruct the classical SIS model by explicitly considering neighbourhood interactions among susceptible and infected individuals (Keeling, 1999; Payne, 2019). We denote by *P*_*S*_ and *P*_*I*_ the probabilities that a randomly chosen individual is susceptible and infected, respectively. These probabilities represent the global frequencies (*S*/*N*) and (*I*/*N*) of susceptible and infected individuals, where *S* and *I* are the number of susceptible and infected individuals in a population of size *N* = *S* + *I*, with *P*_*S*_ + *P*_*I*_ = 1. The disease is transmitted locally when a susceptible individual comes into a direct contact with an infected individual present in its own neighborhood. The pair-approximation method describes the average local frequency of infected individuals present in the neighborhood of a susceptible individual. The number of nearest-neighbouring sites of a susceptible individual is quantified by the positive integer z. The average local frequency of infected individuals in the nearest neighborhoods of a susceptible individual is quantified by *q*_*I*|*S*_, representing the conditional probability that a randomly chosen individual in the neighborhood of a susceptible individual is infected (Harada and Iwasa, 1994). The local frequency of infected individuals is the number of infected individuals in the neighborhood of a susceptible individual, averaged over the number of nearest-neighbouring sites z. A susceptible individual can be infected if at least one of neighbours in the *z* sites is infected.

The disease transmission rate is denoted as β. The infected individual recovers at rate *γ* in a period of time 1/*γ*. In the SIS model, a susceptible individual is assumed to become susceptible again after recovery.

The differential equations governing the global disease dynamics are described by

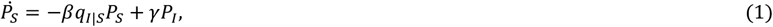

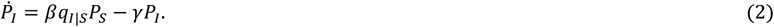

The first term in Eqns. (1) and (2) describes the neighborhood interaction between susceptible and infected individuals. The last term describes the overall recovery rate. The conditional probability *q*_*I*|*S*_ needs to be specified. It is mathematically defined as *q*_*I*|*S*_ = *P*_*SI*_/*P*_*S*_ (Harada and Iwasa, 1994), where the pair *P*_*SI*_ = *P*_*IS*_ represents the joint probability that a randomly chosen neighbouring pair of individuals are susceptible and infected. Notably, there are four possibilities of choosing a neighbouring pair of individuals at random: *P*_*II*_, *P*_*SI*_, *P*_*IS*_, and *P*_*SS*_, with *P*_*II*_ + 2*P*_*SI*_ + *P*_*SS*_ = 1.

Following (Keeling, 1999; Payne, 2019), we construct differential equations governing the dynamics of neighbouring pairs. The following transitions contribute to the dynamics of the *P*_*SI*_ pair:

1. A pair of infected individuals *P*_*II*_ contributes positively to the pair *P*_*SI*_ if one of the infected individuals in the *P*_*II*_ pair has been recovered
2. A pair of susceptible individuals *P*_*SS*_ contributes positively to the pair *P*_*SI*_ if one of the susceptible individuals in the *P*_*SS*_ pair has been infected by one of its direct neighbour from outside the pair, i.e., from the remaining nearest-neighbouring sites (*z* − 1).
3. A pair of susceptible and infected individuals *P*_*SI*_ contributes negatively to the pair *P*_*SI*_ if the susceptible individual in the *P*_*SI*_ pair has been infected by its direct neighbour in the pair
4. A pair of susceptible and infected individuals *P*_*SI*_ contributes negatively to the pair *P*_*SI*_ if the susceptible individual in the *P*_*SI*_ pair has been infected by one of its direct neighbour from outside the pair, i.e., from the remaining nearest-neighbouring sites (*z* − 1).
5. A pair of susceptible and infected individuals *P*_*SI*_ contributes negatively to the pair *P*_*SI*_ if the susceptible individual in the *P*_*SI*_ pair has been recovered

Using the pair-approximation approach (Harada and Iwasa, 1994; Keeling, 1999; Mohammed et al., 2023), the dynamics of the *P*_*SI*_ pair are thus described by

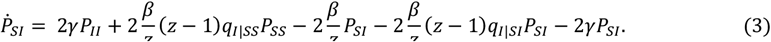

To reduce the model complexity, we replace the neighbouring pair *P*_*II*_ by (1 − *P*_*SS*_ − 2*P*_*SI*_), and we only need to describe the *P*_*SS*_ pair dynamics to construct an equation for the average local frequency *q*_*I*|*S*_. In Eqn. (3), the first term indicates the transition from *P*_*II*_ to either *P*_*SI*_ or *P*_*IS*_, thus the factor 2. In the subsequent term, the quantity β/z represents disease transmission rate per infected individual present in the neighborhood of a susceptible individual, and the transition from *P*_*SS*_ to either *P*_*SI*_ or *P*_*IS*_ will require an infected individual from the remaining neighbouring sites (*z* − 1), thus the multiplication by the conditional probability *q*_*I*|*SS*_. This conditional probability would require the computation of higher-order frequencies, and would not allow to obtain a closed system of differential equations. Therefore, pair-approximation method ignores the effect of the neighbour-of-the neighbour (it assumes correlation is weaker with non-direct neighbours, i.e., correlation between sites decreases exponentially with the distance between them, (see, e.g., Harada and Iwasa, 1994) and approximates *q*_*I*|*SS*_ ≈ *q*_*I*|*S*_ and *q*_*I*|*SI*_ ≈ *q*_*I*|*S*_. That is, correlations with non-nearest neighbours are approximately reconstructed from nearest neighbour correlations. Similar explanations apply to the other terms in Eqn. (3).

The following transitions contribute to the dynamics of the *P*_*SS*_ pair:

1. A pair of susceptible and infected individuals *P*_*SI*_ contributes positively to the pair *P*_*SS*_ if the susceptible individual in the pair *P*_*SI*_ has been recovered
2. A pair of susceptible individuals *P*_*SS*_ contributes negatively to the pair *P*_*SS*_ if any of the susceptible individuals in the pair *P*_*SS*_ has been infected by its direct neighbour from outside the pair, i.e., from the remaining nearest-neighbouring sites (*z* − 1).

The dynamics of the neighbouring pair *P*_*SS*_ are thus described by

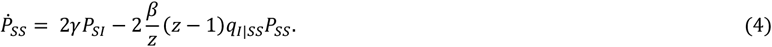

Using the definition of the local frequency of infected individuals *q*_*I*|*S*_ = *P*_*SI*_/*P*_*S*_, and taking the time derivative of both sides, the dynamics of the average local frequency of infected individuals in the neighborhoods of a susceptible individual are described by

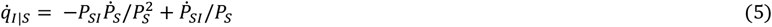

Equations (1) – (5) represent the reconstructed SIS model under neighborhood interactions among susceptible and infected individuals present in the population. Equations (1) and (2) corresponds to the mean-field approximation model when *q*_*I*|*S*_ is replaced by *P*_*I*_.

### 2.2 The basic reproduction number

As defined in the traditional SIS model (Daley and Gani, 2001), the basis reproduction number is given as *R*_0_ = β/*γ*. According to the mean-field SIS model, a disease with *R*_0_ < 1 will die out and persist if *R*_0_ > 1. The bifurcation point occurs at *R*_0_ = 1, separating the extinction and persistence regions of the disease.

### 2.3 Numerical analysis

The numerical simulation of the pair-approximation and the mean-field approximation SIS models has been implemented in MATLAB. Parameter values used to simulate the model are provided in *Table 1*.

**Table 1:**
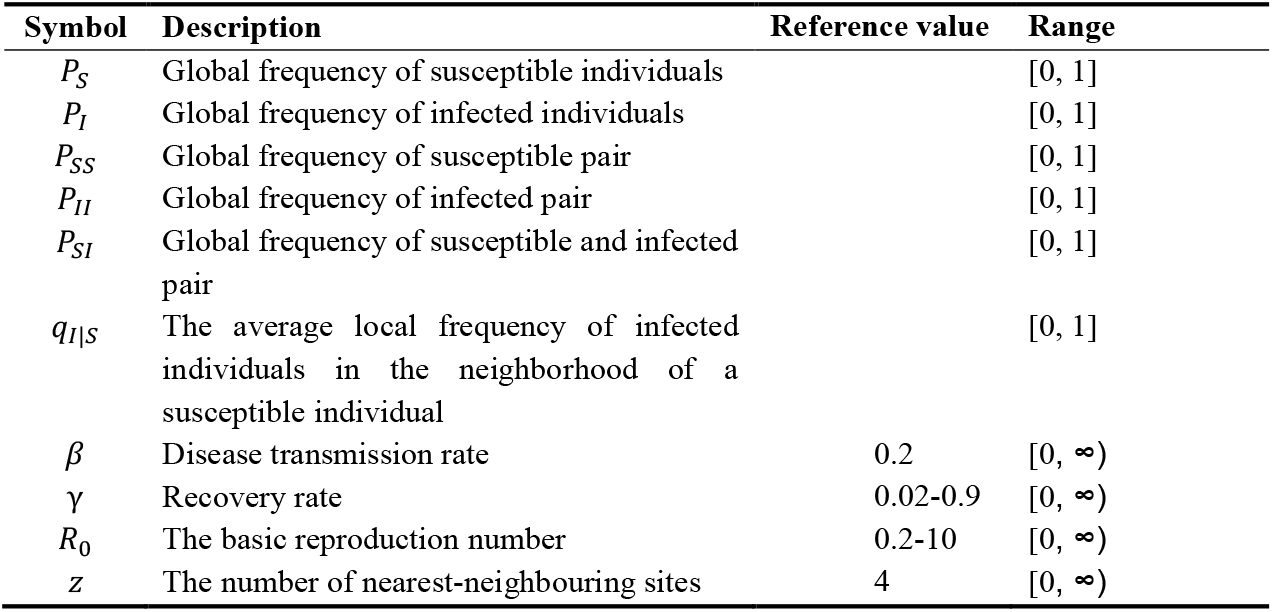
Variable and parameter descriptions, and numerical values used in the model.

## 3 Results

We showcased the SIS model simulation for the global frequencies of susceptible and infected individuals, *P*_*S*_ and *P*_*I*_, respectively. The dynamics of neighbouring pairs (*P*_*SI*_ and *P*_*SS*_) and the average local dynamics of infected individuals present in the neighborhood of a susceptible individual, *q*_*I*|*S*_. We compared results produced by the pair-approximation model with previous predictions by the classical mean-field SIS model.

In the pair-approximation model, the number of nearest-neighbouring sites of a susceptible individual *z* reflects the potential number of susceptible and infected individuals present within the direct neighbourhood of a susceptible individual. In the mean-field approximation model, a susceptible individual has the same likelihood of interacting with all infected individuals present within a population. That is a very unlikely situation and can affect predictions of disease outbreaks. However, it is intuitive to assume that a susceptible individual can typically get infected when interacts with an infected individual present within its direct four neighbouring sites (i.e., *z* = 4).

We first restricted our model to four nearest-neighbouring sites. We found that results produced by the pair-approximation model significantly deviated from predictions by the mean-field approximation. This deviation occurred, particularly, close to the epidemic threshold, *R*_0_ = 2.5 and *R*_0_ = 1.17 (Fig. 1b and c). For large and small values of *R*_0_ (10 and 0.8), corresponding to the sustained transmission and extinction of the disease, the two models produced very similar results (Fig. 1a and d).

**Fig. 1.**
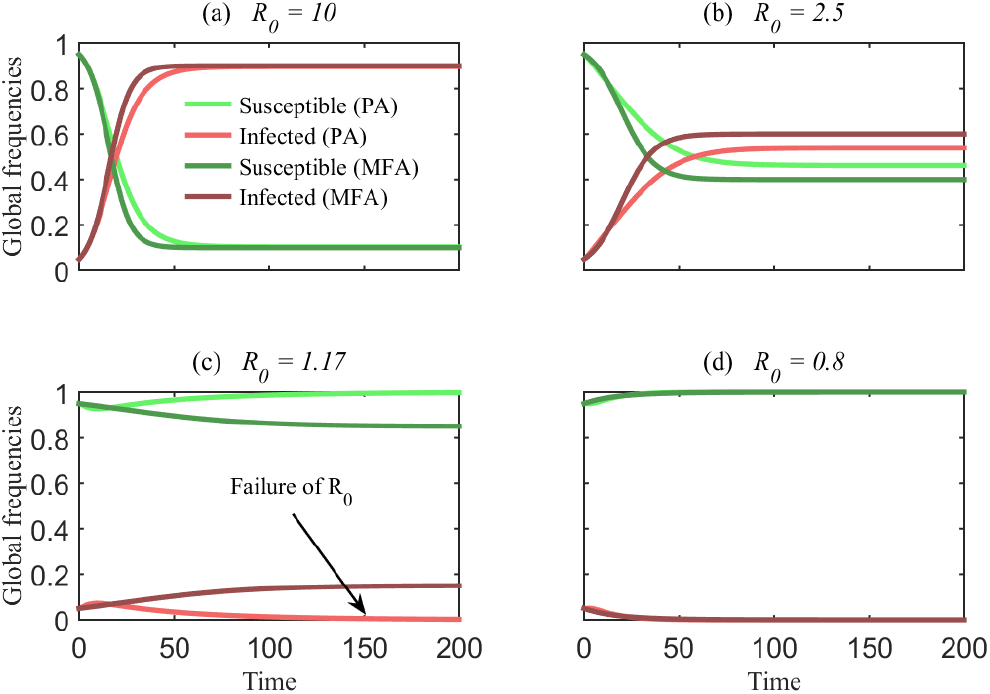
The temporal dynamics of the global frequencies of susceptible (*P*_*S*_) and infected (*P*_*I*_) individuals under four scenarios of the *R*_0_ value (10, 2.5, 1.17, and 0.8). The global frequencies of the pair-approximation (PA) model are represented by the bright-green and red curves, respectively, while the dark curves correspond to the mean-field approximation (MFA) model. The x-axis represents the simulation time, and the y-axis represents the global frequencies. The initial conditions: *P*_*S*_(0) = 0.95, *P*_*I*_(0) = 1 − *P*_*S*_(0), *P*_*SI*_(0) = 0.05, *P*_*SS*_(0) = 0.8, and *q*_*IS*_(0) = *P*_*SI*_(0)/*P*_*S*_(0). Parameters used: β = 0.2, *R*_0_ = 10, 2.5, 1.17, and 0.8, *γ* = β/*R*_0_, and *z* = 4.

According to the classical mean-field SIS model, the disease can spread out and persist when *R*_0_ = β/*γ* > 1, and this principle has been widely accepted for this specific model. However, results of the proposed pair-approximation model demonstrated scenarios where the disease dies out even if *R*_0_ > 1 (Fig. 1c; and Fig. 3a). This divergence is likely driven by the consideration of neighborhood interactions between susceptible and infected individuals in the pair-approximation model. In fact, this is the primary difference between the mean-field and the pair-approximation models presented in this work.

Furthermore, we simulated the dynamics of the reconstructed SIS model for a large number of nearest-neighboring sites (*z* = 100) of a susceptible individual. This indicates that a susceptible individual can potentially interact with 100 individuals in its surroundings, an unlikely situation. Some of these 100 individuals can be infected and can transmit the disease to this susceptible individual at transmission rate β. By considering a very large number of individuals in the neighborhoods of a susceptible individual, the pair-approximation model corresponds to the mean-field approximation model. Therefore, our results demonstrated almost an identical behaviour of the disease outbreak dynamics, produced by the pair-approximation and the mean-field approximation models (Fig. 2; and Fig. 3b). The two models approached the same long-term attractor of the disease, with a small difference in the initial transient phase.

**Fig. 2.**
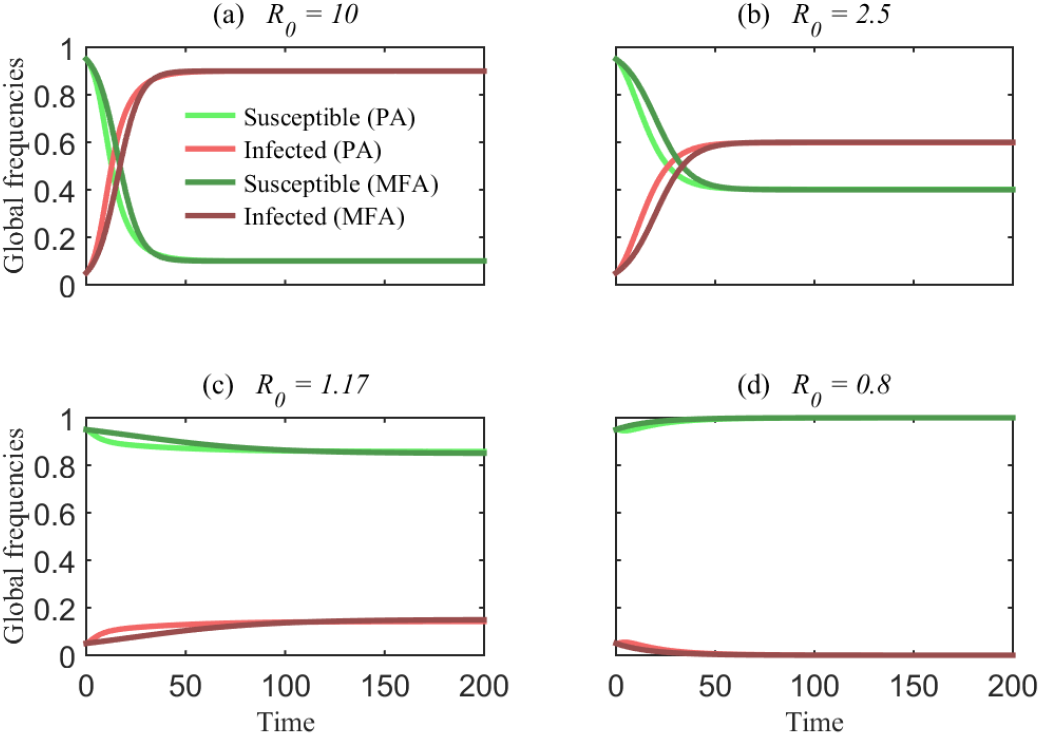
The temporal dynamics of the global frequencies of susceptible (*P*_*S*_) and infected (*P*_*I*_) individuals under four scenarios of the *R*_0_ value (10, 2.5, 1.17, and 0.8). The description, initial conditions, and parameters used are the same as in the caption of figure 1, and *z* = 100.

**Fig. 3.**
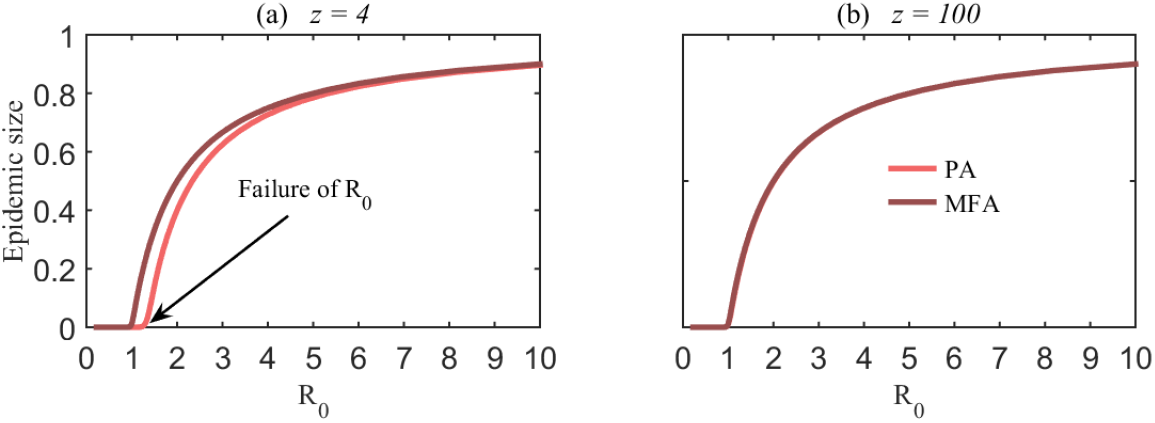
The equilibrium dynamics of the epidemic size (y-axis) as a function the basic reproduction number *R*_0_ (x-axis), predicted by the pair-approximation (PA) model (bright curve) and the mean-field approximation (MFA) model (dark curve). Initial conditions and parameter values are the same as in the caption of figure 1.

We systematically assessed the influence of the basic reproduction number on the epidemic size and compared the corresponding results of the two models. Away from the epidemic threshold (*R*_0_ = 1), the outcome of the disease transmission is intuitively straightforward. Of a particular interest is to predict the disease transmission near the epidemic threshold. Our findings underscored significant differences between the results of the two models (Fig. 3a), especially around the epidemic threshold.

At small to moderate values of *R*_0_, the epidemic size predicted by the pair-approximation model is lower than that predicted by the mean-field approximation model (Fig. 3a). More precisely, the epidemic size curve produced by the pair-approximation model is shifted forward, extending the disease extinction region. Notably, this forward shift emerged only under the consideration of four nearest-neighbouring individuals (*z* = 4). The results of the two models produced the exact dependence of the epidemic size on the basic reproduction number *R*_0_ when the number of nearest-neighbouring sites is chosen to be very large, *z* = 100 (Fig. 3b).

The temporal dynamics for the average local frequency of infected individuals within the neighbourhood a susceptible individual, and the dynamics of neighbouring pairs are shown for a number of nearest-neighbouring sites *z* = 4 (Fig. 4) and *z* = 100 (Fig. 5).

**Fig. 4.**
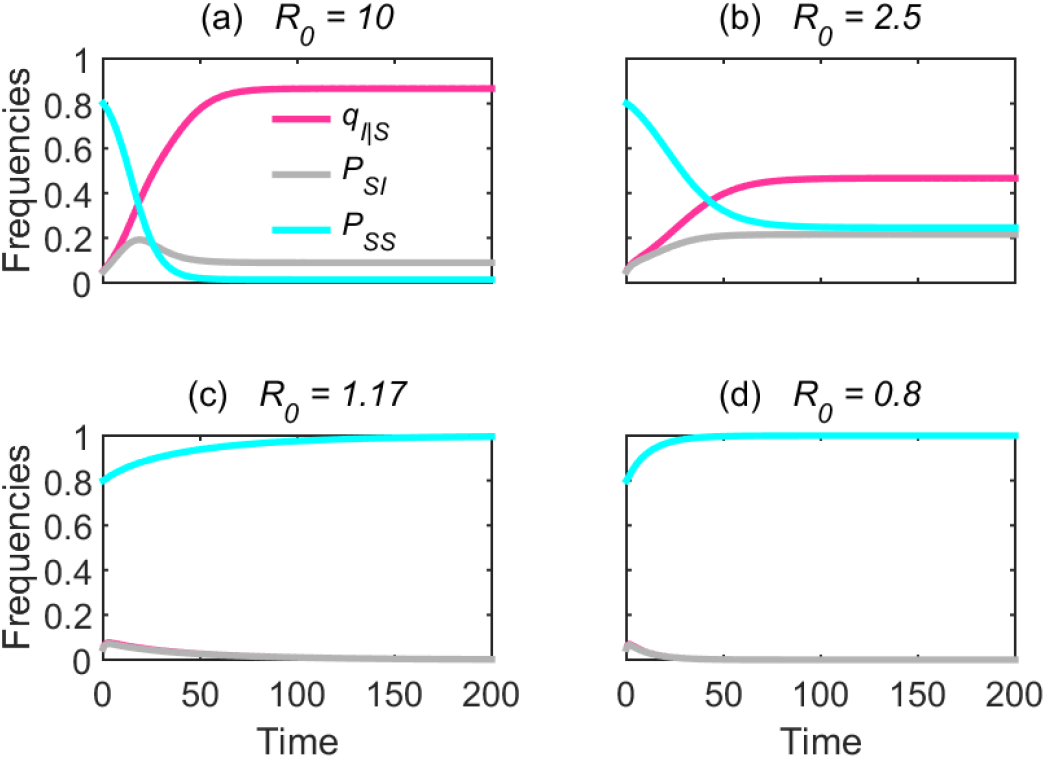
The temporal dynamics of the average local frequency (*q*_*I*|*S*_, pink curve), neighbouring pairs of susceptible and infected individuals (*P*_*SI*_, grey curve), and neighbouring pairs of susceptible individuals (*P*_*SS*_, cyan curve) predicted by the pair-approximation model under four scenarios of the *R*_0_ value (10, 2.5, 1.17, and 0.8). The x-axis represents the simulation time, and the y-axis represents the frequencies. The initial conditions: *P*_*S*_(0) = 0.95, *P*_*I*_(0) = 1 − *P*_*S*_(0), *P*_*SI*_(0) = 0.05, *P*_*SS*_(0) = 0.8, and *q*_*IS*_(0) = *P*_*SI*_(0)/*P*_*S*_(0). Parameters used: β = 0.2, *R*_0_ = 10, 2.5, 1.17, and 0.8, *γ* = β/*R*_0_, and *z* = 4.

**Fig. 5.**
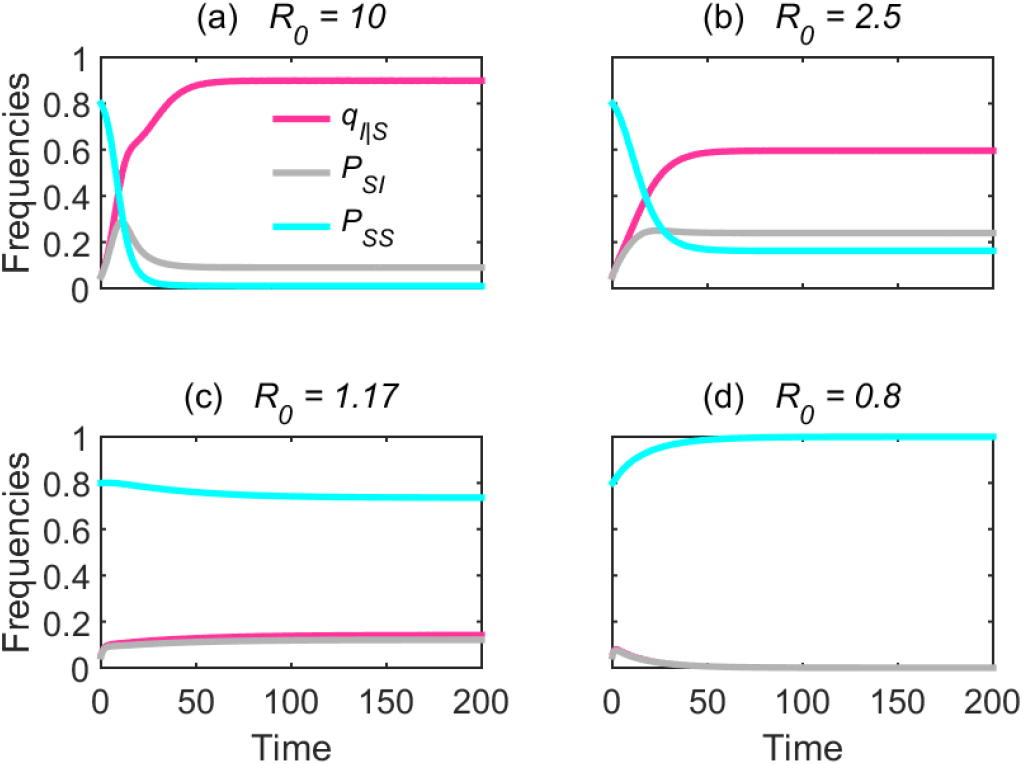
The temporal dynamics of the average local frequency (*q*_*I*|*S*_, pink curve), neighbouring pairs of susceptible and infected individuals (*P*_*SI*_, grey curve), and neighbouring pairs of susceptible individuals (*P*_*SS*_, cyan curve) predicted by the pair-approximation model under four scenarios of the *R*_0_ value (10, 2.5, 1.17, and 0.8). Initial conditions and parameter values are the same as in the caption of figure 4.

Results demonstrated that the average local frequency of infected individuals decreases as the basic reproduction number *R*_0_ increases (Fig. 4 and 5). A small number of infected individuals are present in the neighbourhoods of each susceptible individual when the basic reproduction number is small. Thus, the probability of choosing an infected individual from the neighbourhood of a susceptible individual is low. The average local frequency of infected individuals will also reflect the probability of choosing a neighbouring pair of susceptible and infected individuals. The probability of randomly choosing a neighbouring pair of susceptible and infected *P*_*SI*_ depends on the absolute difference between the global probabilities *P*_*S*_ and *P*_*I*_. The probability of choosing the pair *P*_*SI*_ is small when the absolute difference is high, and vice versa. The probability of randomly choosing the neighbouring pair *P*_*SS*_ typically increases as the global frequency *P*_*S*_ increases. Thus, a small values of *R*_0_ will lead to higher probabilities of a neighbouring pair of susceptible individuals (Fig. 4). However, under the consideration of very large number of nearest-neighbouring sites, the probability of choosing an infected individual from the neighbourhood of a susceptible individual is relatively high when *R*_0_ is slightly greater than one (Fig. 5c).

## 4 Conclusion

The basic reproduction number *R*_0_ has been used for several decades as an indicator of whether the disease will die out or persist. It is widely acknowledged that the disease is likely to die out if *R*_0_ < 1, while persist if *R*_0_ > 1. In this work, we reconstructed the classical SIS model using the pair-approximation method (Harada and Iwasa, 1994; Keeling, 1999; Payne, 2019), and presented a theoretical evidence that a disease with *R*_0_ > 1 dies out in a simple SIS model. Despite this simplicity of the model, results produced by the pair-approximation model deviated pronouncedly from predictions by the mean-field approximation model. The advantage of the pair-approximation approach over the commonly-used mean-field approximation is that it explicitly accounts for neighborhood interactions among susceptible and infected individuals. However, a recent pair-approximation SIS model produced results in alignment with predictions by the classical mean-field SIS model, yet diverging from our findings in terms of the dependence of epidemic size on *R*_0_ (Payne, 2019). That model, while informative, solely focused on dynamics of pairs and did not consider the average local frequency dynamics of infected individuals; thus, overlooked the effect of the number of nearest-neighboring sites of a susceptible individual on the disease transmission dynamics. In the realm of more intricate models of infectious diseases, it remains an open question to what extent mean-field approximation predictions deviate from predictions produced by models that consider neighborhood interactions (Keeling, 1999). We suggest a crucial need for redefining the basic reproduction number on a smaller spatial scale and taking the average *R*_0_ over a global scale, rather than applying it globally to an entire population. This redefinition may depend on the average number of infected individuals within the neighbourhoods of a susceptible individual (Keeling, 1999). This fundamental epidemiological concept of *R*_0_ might otherwise be misleading (Li et al., 2011).

## Data availability

This study does not include or use data.

